# Mapping the Cellular Etiology of Schizophrenia and Diverse Brain Phenotypes

**DOI:** 10.1101/2024.10.21.24315695

**Authors:** Laramie E Duncan, Tayden Li, Madeleine Salem, Will Li, Leili Mortazavi, Hazal Senturk, Naghmeh Shargh, Sam Vesuna, Hanyang Shen, Jong Yoon, Gordon Wang, Jacob Ballon, Longzhi Tan, Brandon Scott Pruett, Brian Knutson, Karl Deisseroth, William J Giardino

## Abstract

Psychiatric disorders account for a substantial fraction of the world’s disease burden^1^, and yet the development of novel therapeutics has been notoriously slow^2^. Likely contributing factors include the complexity of the human brain and the high polygenicity of psychiatric disorders^3–5^, meaning that thousands of genetic factors contribute to disease risk. Fortunately, technological advances have enabled comprehensive surveys of human brain cell types using transcriptomes from single nuclei (snRNAseq)^6–8^. Additionally, genome-wide association studies (GWAS) have linked thousands of risk loci to psychiatric disorders^9–11^. Here, we combined these two landmark data resources to infer the cell types involved in the etiology of schizophrenia and comparison phenotypes. This work demonstrated: 1) cell types that are concordant with prior findings about schizophrenia, 2) novel cell type associations for schizophrenia, 3) greater molecular specificity regarding schizophrenia-associated cell types than was previously available, 4) evidence that well powered genome-wide and brain-wide datasets are required for these analyses, 5) distinct cellular profiles for five brain-related phenotypes, 6) a prototype for a cell-type based classification system for psychiatric and other brain disorders, and 7) a roadmap toward drug repurposing, novel drug development, and personalized treatment recommendations. Thus, this work formalizes a data-driven, cellular and molecular model of complex brain disorders.

## Introduction

Approximately one in five adults has a psychiatric disorder^12^. While these disorders may resolve on their own or with treatment, for many, they represent lifelong afflictions.

Schizophrenia is often considered the cardinal adult psychiatric disorder given its severity, chronicity, and enduring worldwide distribution^13–15^. Like all other major psychiatric disorders (depression, substance use disorders, PTSD, etc.), the etiology of schizophrenia depends both on genetic and environmental factors. All such disorders are moderately to highly heritable, and schizophrenia is one of the most heritable of these “complex polygenic” disorders, with heritability estimates from twin studies of approximately 80%^16,17^.

Accordingly, investment in research on schizophrenia has successfully generated knowledge about its genetic basis. Massive human genome-wide association studies (GWAS) encompassing millions of individuals, often led by the international Psychiatric Genomics Consortium (PGC), showed that common genetic variants account for a substantial fraction of population liability to schizophrenia as well as other psychiatric disorders^3,9,18–20^. For instance, the most recent GWAS of schizophrenia in a sample of 320,404 participants found 287 risk loci across the human genome (see **Figure 1A**). These loci exceeded a stringent, international threshold for statistical significance (*p*<5x10^-8^), which corrects for tests of multiple hypotheses. While these landmark studies have revealed novel discoveries about psychiatric disorders, they also raised a new challenge: determining the physiological relevance of associated genomic loci.

**Figure 1.**
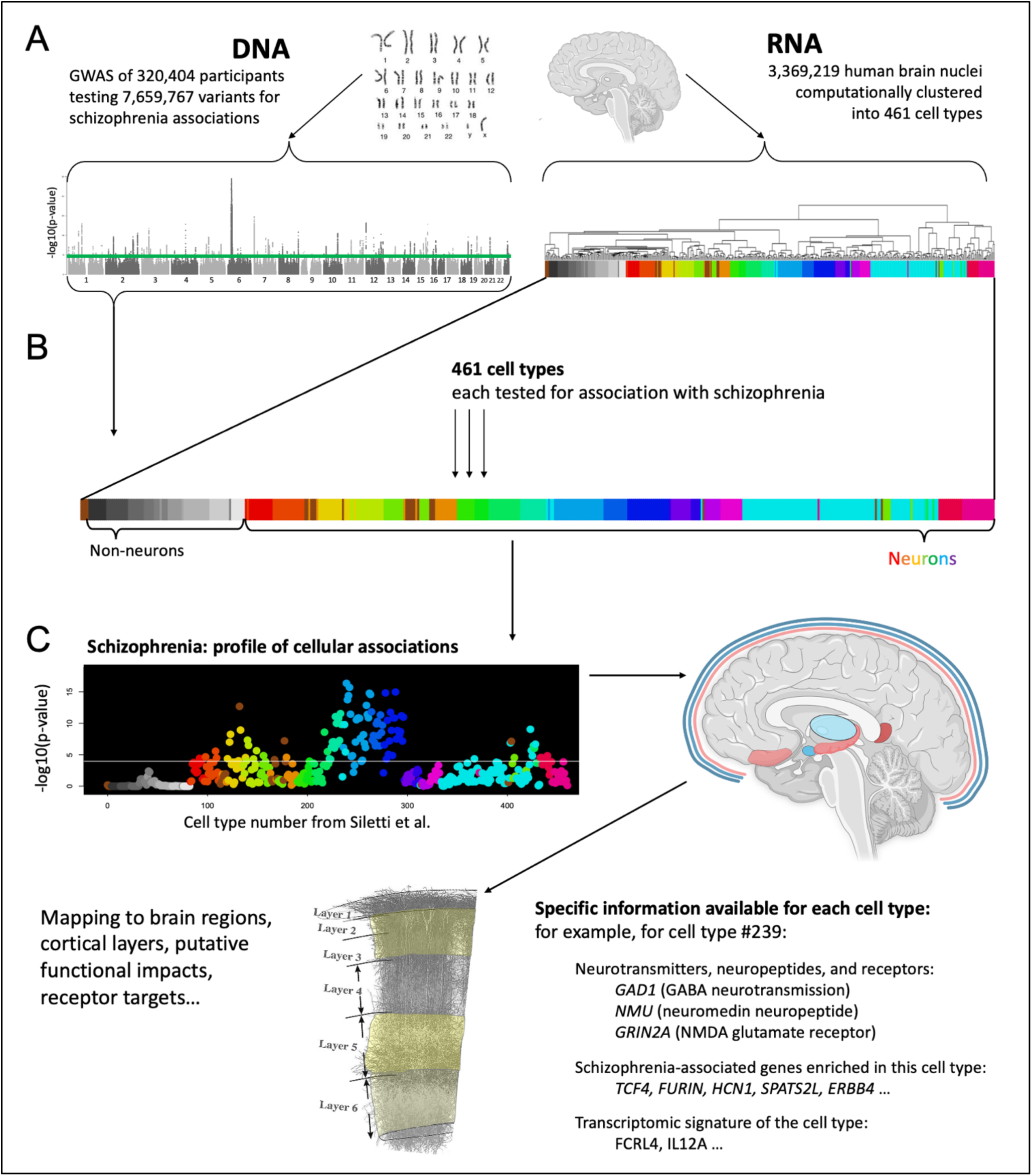
Approach for systematically testing 461 human brain cell types for association with schizophrenia. We tested whether genes associated with schizophrenia were preferentially expressed in one or more brain cell types. **A.** We used two types of human genome-wide data. **Left**: Genome wide association study (GWAS) results from the most recent (2022) schizophrenia GWAS which identified 287 loci for schizophrenia in a sample of 320,404 participants. GWAS results are depicted in a Manhattan plot with the chromosomal position on the x-axis and statistical significance (-log10(p)) on the y-axis, for each genetic variant tested. **Right**: cell type data from the most comprehensive human brain snRNAseq study to date (3,369,219 cells from 106 brain regions, clustered into 461 statistical clusters, referred to here as cell types). For the 461 cell types, colors reflect superclusters as assigned in Siletti et al. Here we used grayscale to denote non-neuronal cell types and rainbow colors for neuronal cell types (see Figure 2 for complete color coding). **B.** Each of the 461 cell types was tested for association with schizophrenia. As detailed in the methods, we first calculated ‘specificity’ scores using established procedures to quantify the fraction of each gene’s total expression that was found in each cell type (for 16,641 genes, see Methods). Specificity values ranged from 0-1, and for each gene the specificity scores summed to 1 over the 461 cell types (by definition). MAGMA was first used to quantify schizophrenia associations for each gene (as applied to GWAS data). Next, for each of the 461 cell types (analyzed separately), we used regression (implemented in MAGMA) to test for linear relationships between specificity scores and schizophrenia associations of those genes, while correcting for known potential confounding variables. **C.** Results are depicted first as a cell type profiles (here for schizophrenia), then associated cell types are described with functionally relevant details (e.g., brain structure, neurotransmitters, receptors, cortical layer localization).

Polygenic influences on schizophrenia offer a powerful entry point for discovering disease etiology, when combined with data about how genes are used in specific brain cell types. Gene expression from a common nuclear genome is largely what generates cellular diversity in the body, and newly available technology allows measurement of gene expression (RNA molecule counts) from individual cells or the nuclei within those cells via single nucleus RNA sequencing (snRNAseq^i^). Moreover, computational clustering of cellular transcriptomes in snRNAseq datasets readily reveals known as well as novel cell types, and consequently this approach is being used to create comprehensive atlases of cell types in human and animal organs^10,21,22^. Recently, a landmark snRNAseq dataset for the human brain was released^6^. Previous human studies had surveyed only small numbers of brain regions. Thus, Siletti et al.’s^6^ analysis of 106 brain regions and 3,369,219 individual nuclei is by far the most comprehensive human snRNAseq dataset available to date. This dataset afforded statistical clustering of 461 cell types in normal human brains, 378 of which are neuronal. The right side of **Figure 1A**, adapted from Siletti et al., shows these 461 cell types as the ends of the leaves of a dendrogram. Using this critical dataset which specifies gene expression in hundreds of healthy, adult human brain cell types, we were able to map the polygenic risk for schizophrenia^9^ (as quantified in schizophrenia GWAS) to specific human brain cell types and their anatomical locations. This was accomplished in an entirely data-driven manner using the leading available genome-wide and brain-wide datasets and is therefore “unbiased” with respect to researcher-driven hypotheses about which cell types should be associated with schizophrenia.

### Present study

Based on the work of Bryois et al., Watanabe et al., and Skene et al. (who used rodent data and more limited human snRNAseq datasets^23–25^ available at the time), we used MAGMA^26,27^ software for analysis of GWAS data to analyze the schizophrenia GWAS results implementing gene property analysis. We determined the cell types that had gene usage that was positively correlated with schizophrenia gene associations, while adjusting for potential confounders (e.g. gene size, gene density; see methods for details). We then used conditional analyses to empirically select representative cell types among all significant results for further discussion.

The 461 cell types tested here are hierarchically organized and numerically close cell types tend to have similar gene expression profiles. This means that there are not 461 statistically independent cell types and that Bonferroni correction is overly stringent. Nevertheless, we implemented Bonferroni correction to be conservative. This also means that significant cell types may be significant for two reasons: either they are driving a phenotypic association (truly associated), or they appear associated because they share overlapping gene expression with a cell type (or types) driving associations. The latter scenario (correlated statistical tests) is a common problem relevant to diverse domains of research, and both statistical and experimental procedures can clarify truly associated cell types. Here, we used conditional analysis (see Methods), the leading statistical approach for handling this issue in this type of analysis^24,28^. In brief, each cell type was tested for association while controlling for each of the other cell types. Interpretation-wise, it is most conservative to only focus on these ‘significant independent’ cell types, hence our primary focus on the significant independent cell types in this manuscript.

Summarizing, we used robust statistical procedures to combine two massive, unbiased, genome-wide and brain-wide datasets to systematically test which brain cell types were linked with schizophrenia. To validate this approach and determine disease-specificity, we also analyzed four comparison phenotypes (alcohol consumed per week^29^, sleep duration per night^11^, multiple sclerosis^30^, and Alzheimer’s disease^31^). Results from these analyses matched prior expectations (e.g. of immune cell relevance to multiple sclerosis) and also revealed novel cell type associations. In sum, these findings yielded a data driven map of the cellular etiology of schizophrenia, demonstrated clear extensibility to other brain phenotypes, and suggested a tractable roadmap toward personalized psychiatric treatment using low-cost genotype data.

Throughout this manuscript, please refer to **Supplementary Table S1** for more complete information about the 461 cell types analyzed in this report (numbered 0-460, per Siletti et al.^6^).

### Schizophrenia associated cell types

Of the 109 total cell types that were significantly associated with schizophrenia, there were ten significant independent cell types (see **Figure 2**), as follows. The most significant cell type was a subtype of somatostatin interneurons (#239, *p*=4.3x10^-17^). The next two significant independent cell types were also cortical: PAX6 interneurons distributed widely across the cortex (#278, *p*=1.5x10^-15^) and an excitatory cell type found almost exclusively in retrosplenial cortex (#132, *p*=2.1x10^-13^, 91% of cells from retrosplenial cortex). Note that the PAX6 interneurons (#239) were annotated as GABA/VGLUT3, indicating co-expression of GABA and glutamate, which is relatively uncommon. Fourth and fifth were two distinct inhibitory amygdala neuron types #233 (*p*=2.8x10^-12^) and #423 (*p*=9.0x10^-10^), see below for more details. The remaining five cell types had neurons primarily from the prefrontal cortex (#404, *p*=7.3x10^-8^, specifically from Broadmann area 14), thalamus (#440, *p*=1.4x10^-5^), cortex-wide excitatory neurons annotated to deep layer 6b (#98, *p*=2.2x10^-5^), and two excitatory hippocampal neuron types (#179 *p*=2.6x10^-5^ and #202 *p*=1.1x10^-4^). In addition to these ten significant independent cell types, other notable significant cell types were medium spiny neurons in the striatum (i.e., caudate and putamen, #222), cortex-wide excitatory neurons in layer 2/3 (#123), as well as cell types preferentially located in visual cortex (#133), septal nuclei (#428), superior colliculus (#433, #367), and substantia innominata (#232).

**Figure 2.**
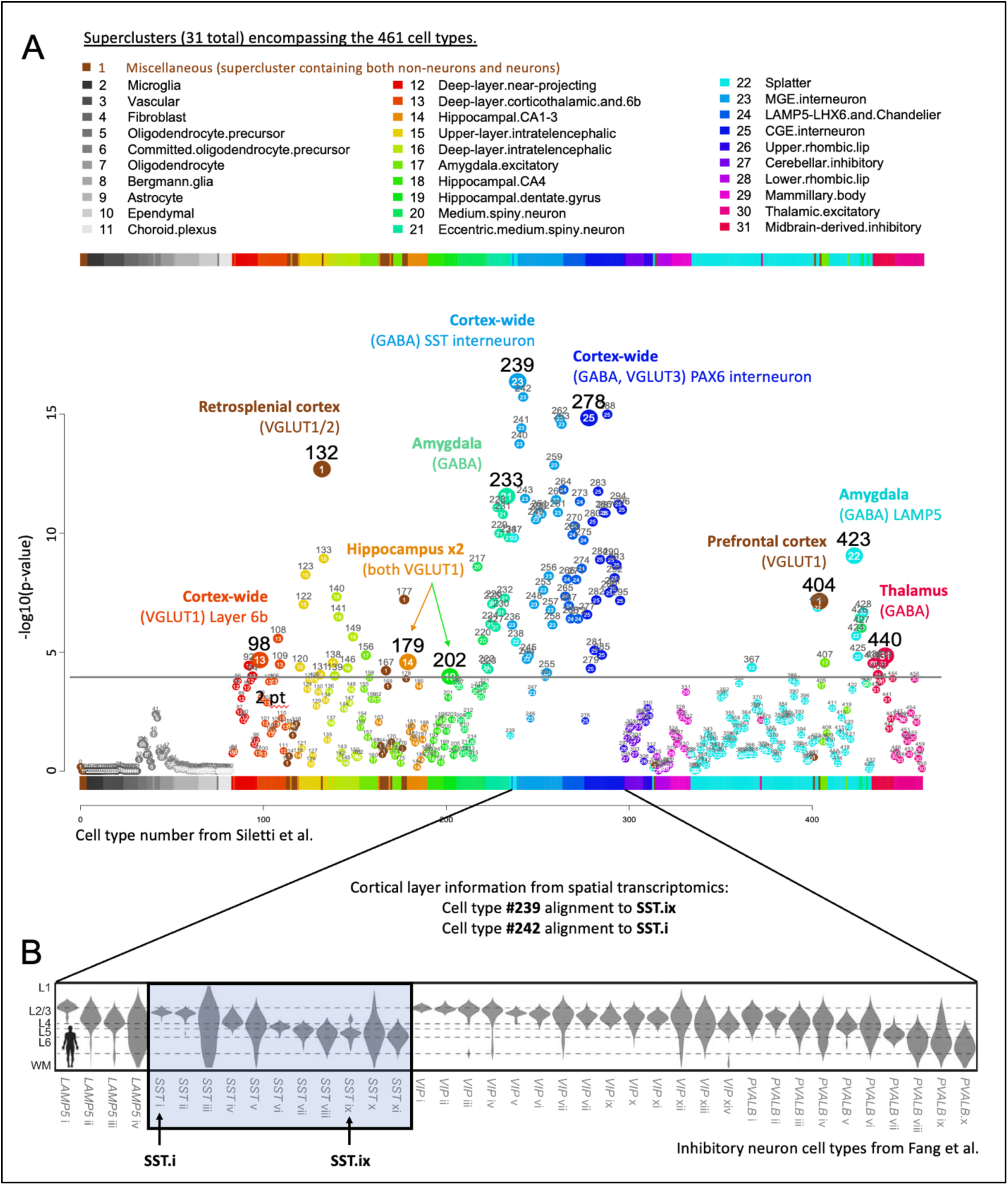
Cell type associations for schizophrenia reveal known and novel cell types for schizophrenia. **A.** Schizophrenia results for 461 cell types are depicted in a scatterplot, with cell type number on the x-axis and statistical significance on the y-axis (-log10(*p*)), such that higher values are more statistically significant). The horizontal gray line denotes Bonferroni correction for the 461 cell types (*p*<.0001). Numbers above points are cell type numbers and numbers within points are supercluster numbers. Larger points denote the ten cell types that were ‘significant independent’ cell types after conditional analyses (see Methods). **B.** The top cell type for schizophrenia (#239) aligned to somatostatin interneuron subtype SST.ix (from Fang et al.) for which cortical layer localization had been determined using spatial transcriptomics. Thus, we inferred that cell type #239 was likely most abundant in cortical layer 5, but also found in cortical layers 6 and 2/3, whereas cell type #242 was likely most abundant in layers 2/3.

### Interneurons

For schizophrenia, arguably the most widely replicated postmortem tissue finding involves aberrant inhibitory neurons in the cortex (interneurons)^32,33^. While both the parvalbumin and somatostatin (SST) subclasses of interneurons have been implicated, the most recent evidence shows the greatest abnormalities in somatostatin interneurons^32,33^. Thus, the present report of the strongest association with somatostatin (SST) interneurons, and many significantly associated interneurons (including parvalbumin), is concordant with leading prior findings. We extended these findings by specifying more subtle subtypes of somatostatin interneurons and by determining the probable cortical layers for these schizophrenia-associated somatostatin interneuron subtypes (#239 and #242^ii^) using additional snRNAseq and spatial transcriptomics datasets.

Specifically, recent studies have identified far more interneuron subtypes than were distinguishable using traditional markers. These transcriptomically-defined cell types reside preferentially in specific cortical layers in both humans and mice^7^. In **Figure 2B**, with a plot adapted from Fang et al.^7^, the cortical layer distribution of human interneuron subtypes is shown (i.e. cortical layers 1-6). We aligned somatostatin subtypes from the Linnarsson^6^ and Zhuang^7^ laboratories, and found that #239 and #242 (from Siletti et al.) best matched SST.ix and SST.i, respectively (from Fang et al.), and that these cell types differed in cortical layer distribution. #239 somatostatin interneurons were most likely to be localized to cortical layer 5 (and less so to layers 6 and 2/3), while #242 somatostatin interneurons were most likely to be in layers 2/3. Alignment to two mouse datasets^8,34^ yielded similar cortical layer localization results, providing additional support for putative layer localizations. Contextualizing these findings, we note that schizophrenia postmortem tissue studies have primarily identified deficits in upper cortical layers, particularly deep layer 3^35,36^, which may be consistent with the primary localization of cell type #242 and, to a lesser extent, #239. Given known disruptions in sensory processing and integration in schizophrenia, it is conceivable that normal cortical layer 5 integration and output functions^37^ are disrupted in schizophrenia due to abnormalities in layer 5 somatostatin interneurons, of the #239 type.

### Amygdala

Within the amygdala, which is a structure known to have diminished volume in schizophrenia^38^, we found 17 significant cell types (including both inhibitory and excitatory), two of which were independent significant cell types (#233 and #423, both inhibitory). Cell type #233 was annotated by Siletti et al as a medium spiny neuron of the “eccentric” subtype, a recently discovered subtype of medium spiny neurons^10^. This general class of neurons – medium spiny neurons – has been frequently linked to schizophrenia, and medium spiny neurons are the dominant inhibitory neuron type of the striatum (caudate and putamen). In addition to the two major subtypes of medium spiny neurons, which are well characterized based on their differential expression of dopamine receptors (D1 versus D2), recent transcriptomic studies made clear that a third subtype of medium spiny neurons also exists. These ‘eccentric’ medium spiny neurons had evaded detection because classical D1/D2 markers do not reliably differentiate the newly named ‘eccentric’ medium spiny neurons^10^. Thus, the linking of this eccentric medium spiny neuron type (#233) to schizophrenia demonstrates multiple benefits of large-scale transcriptomic studies and the unbiased approaches used here to link cell types to schizophrenia. First, scRNAseq studies made possible the detection of the relatively rare (∼4%) but clearly transcriptomically distinct “eccentric” medium spiny neuron subtype^10^.

Second, the brain-wide Siletti et al snRNAseq dataset used here shows that neurons that transcriptomically resemble all three subtypes of medium spiny neurons (D1, D2, and eccentric) are also found outside of the striatum. And third, this approach links these newly discovered, relatively rare, and extra-striatal eccentric medium spiny neurons to schizophrenia. Such a discovery would not have been possible without these large, comprehensive, and “unbiased” datasets that allow us to see more clearly the gaps in prior knowledge, and to discover which newly filled gaps might also provide critical information about schizophrenia etiology^iii^.

The second significant independent amygdala cell type (#423) is inhibitory and while found predominately in amygdala (51% of cells), is also present in the thalamus (20%), hypothalamus (15%), and other brain regions. Notable gene enrichments for this cell type include a cytochrome p450 gene (*CYP19A1*) responsible for the biosynthesis of estrogen, a sodium channel subunit gene (*SCN5A*), a GABA receptor A subunit gene (*GABRQ*), and neurotensin, a neuropeptide that modulates dopamine and other neurotransmitters relevant to psychiatric disorders, which has been investigated as a therapeutic target for schizophrenia^39,40^. Other amygdala cell types associated with schizophrenia, but not deemed independent after conditional analyses, include medium spiny neurons of the D1 (e.g. #220) and D2 (#217) subtypes.

### Hippocampus

Our hippocampal findings provide molecular details that can augment and extend schizophrenia imaging and postmortem tissue findings. Prior work showed lower average volume of the hippocampus and smaller individual hippocampal neurons, as reported in meta-analyses^41^. Here, we have identified specific neuron types that may underlie these hippocampal volume reductions in schizophrenia. Of the 86 neuron types located primarily in the hippocampus (from Siletti et al.’s data^6^), seven were significantly associated with schizophrenia in our analyses. Two of those seven significant hippocampal neuron types were significant independent cell types after conditional analyses (#179 and #202). Cell type #179 is an excitatory neuron type (VGLUT1) and contributing cells came from all major subregions of the hippocampus (subiculum, CA1-4, dentate gyrus). Cell type #202 is also excitatory (VGLUT1) and contributing dissections encompass all major hippocampal subregions except the subiculum. The present report may also be concordant with prior reports of increased hippocampal excitatory (glutamate) neurotransmitter metabolites in individuals with schizophrenia^42^, and based on these results future studies can target more specific subtypes of neurons.

In sum, the primary anatomical locations for the significant independent schizophrenia cell types were three widely distributed across the cortex, two from specific cortical regions (retrosplenial and prefrontal), two from amygdala, two from hippocampus, and one from the thalamus. Of note, the three subcortical structures linked to schizophrenia here (amygdala, hippocampus, and thalamus) are precisely the same subcortical structures found to have the largest volume decreases in schizophrenia patients as compared to controls (Hedges *g* of -.66, -.46, and -.31, respectively) in a recent meta-analysis of brain volume studies in first-episode psychosis^43^. Thus, two completely independent, brain-wide, data-driven approaches to understanding schizophrenia – one using imaging data^43^ and the present approach using genetic data – pointed to the same three subcortical regions: amygdala, hippocampus, and thalamus. In the next section we consider the cell types and brain regions linked to four other phenotypes using this approach.

### Contrasting five brain phenotypes

Prior investigations across different branches of medicine have linked cell types to phenotypes using gene expression and GWAS data^23–25,44^. We sought to establish whether this new human snRNAseq dataset could also afford accurate identification of cell types linked to comparison brain-related phenotypes (alcohol consumed per week, sleep duration per night, multiple sclerosis, and Alzheimer’s disease; see the Methods for the rationale for selecting these phenotypes). As shown in **Figure 3**, expected cell type phenotype pairs were found and are discussed below (see **Supplementary Tables 2-5** for full results).

**Figure 3.**
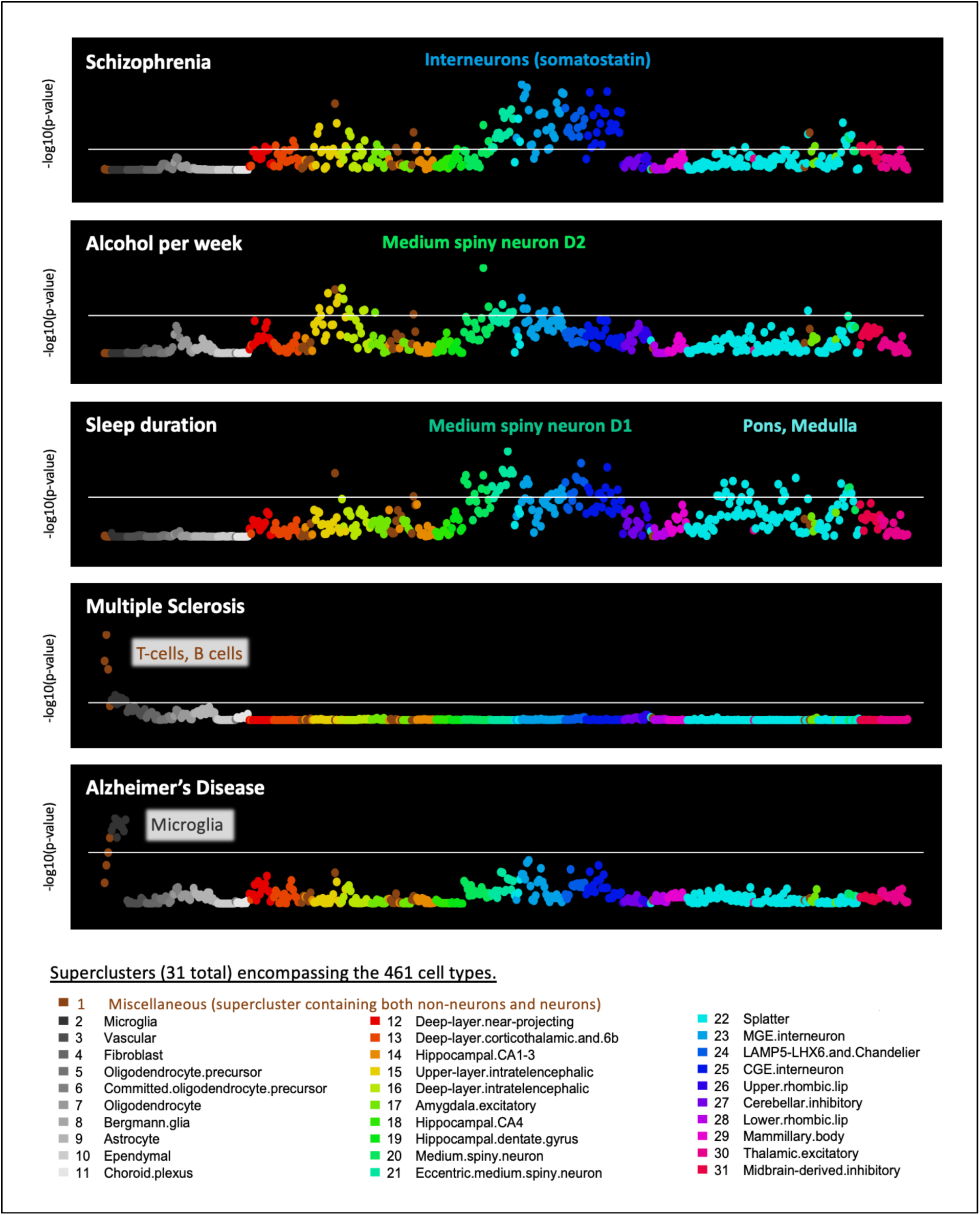
Distinct cell type profiles for five phenotypes. Cell-type associations for five brain-related phenotypes are depicted in scatterplots, with cell type number (0-460) on the x-axis and statistical significance on the y-axis (reported as -log10(*p*), such that higher values are more statistically significant). Horizonal white lines reflect correction for 461 tests (i.e. *p*<0.0001). The first three phenotypes (schizophrenia, alcohol consumed per week, and sleep duration per night) yielded neuronal cell-type associations^iv^. The two neurological phenotypes (multiple sclerosis & Alzheimer’s disease) yielded non-neuronal cell type associations of immune and microglial cell types, respectively. See **Supplementary** Figures 1-4 for larger annotated scatterplots for the comparison phenotypes.

### Schizophrenia, Alcohol consumed per week, and Sleep duration per night

We discussed previously the notability of the somatostatin interneuron associations with schizophrenia (e.g., #239), given prior findings of somatostatin interneuron abnormalities in schizophrenia^32,33^. For alcohol consumed per week, the top cell type was a D2 medium spiny neuron (#217, *p*=1.3x10^-9^), which also matched prior expectations given that D2 medium spiny neurons causally influence alcohol consumption^45–49^. For sleep duration per night, the top cell type was also a medium spiny neuron, but of the D1 type (#231, *p*=2.8x10^-9^). This finding may be consistent with recent findings linking D1-MSNs and sleep, particularly rapid eye movement (REM) sleep^50^. For sleep duration per night, we also highlight associated cell types from the pons (#396, *p*=1.3x10^-6^) and medulla (#386, *p*=5.9x10^-6^) since these structures are key nodes of sleep regulatory circuits^51^. Note that these pons and medulla cell types were not associated with schizophrenia or alcohol consumption, but rather they are specific among these phenotypes for sleep. Note as well that the sleep phenotype had many associations with the newly named “Splatter” supercluster of neurons (described in Siletti et al.^6^), which includes a wide variety of subcortical neuron types. See **Figure 4** for a comparison of the brain regions harboring the significant independent cell types for these three psychiatrically relevant phenotypes.

**Figure 4.**
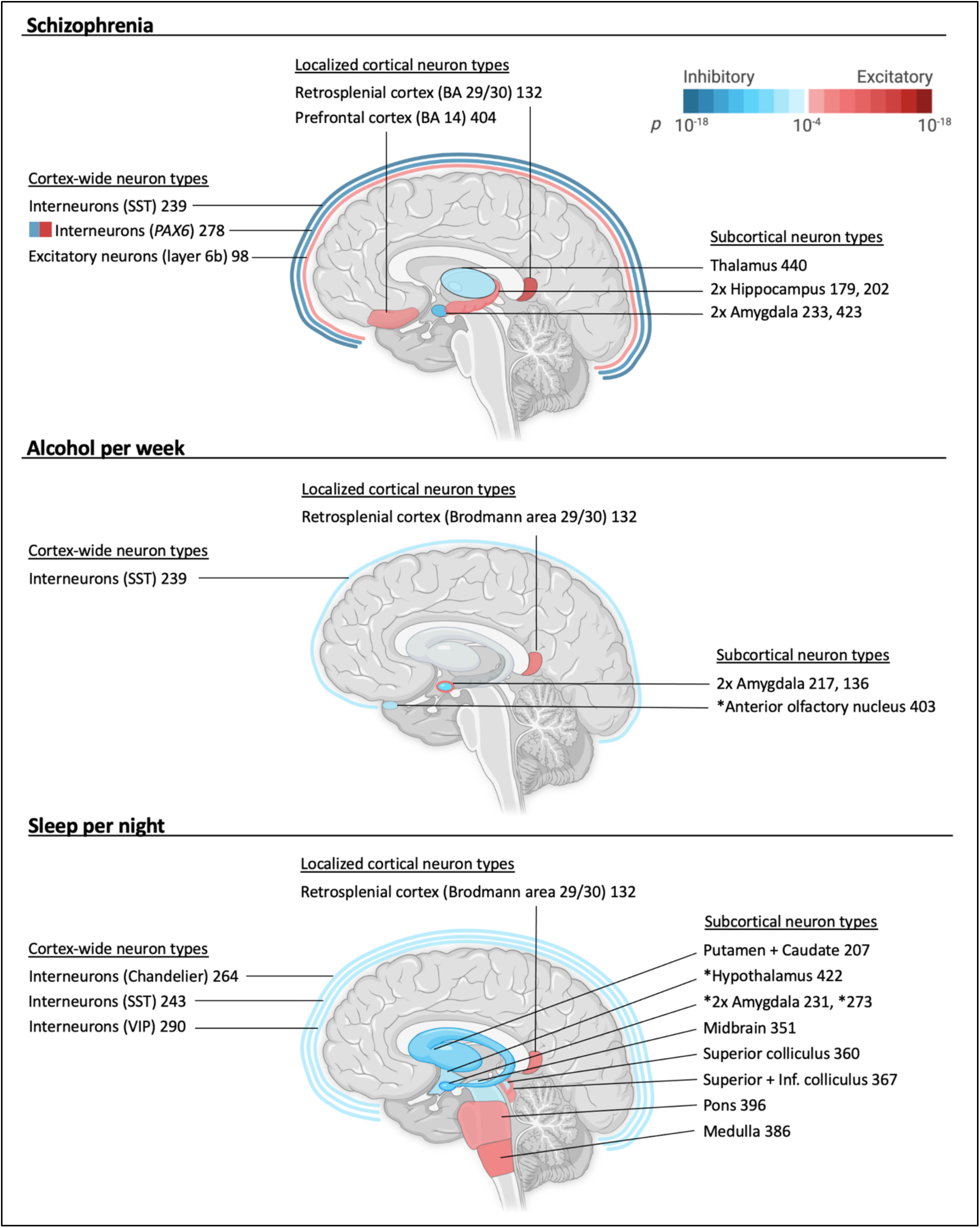
Brain locales of origin for the independent significant cell types associated with schizophrenia, alcohol consumed per week, and sleep duration per night. Inhibitory cell types are depicted in blue, and excitatory cell types are in red, with saturation of color denoting statistical significance. Cell type numbers are given after brief descriptions. Unless an asterisk is present, the regions depicted were the source of >50% (and usually much >50%) of cells for a particular transcriptomic cell type. If there are two excitatory or two inhibitory cell types for a single brain structure, color saturation corresponds to the more significant cell type. If both excitatory and inhibitory cell types are associated with a single structure, then the fill of the structure denotes the more significant association (as opposed to the border). For example, there are two amygdala cell types associated with alcohol per week and the inhibitory cell type #217 is more significant (hence the blue/inhibitory fill). BA=Broadmann area, SST=somatostatin, *PAX6*=paired box 6 gene, VIP=vasoactive intestinal peptide, Inf=inferior. Cell type #278 is annotated as GABA/VGLUT3.

### Multiple sclerosis and Alzheimer’s disease

For multiple sclerosis, an autoimmune condition, the strongest association was a T cell type (cell type #1, *p*=6.0x10^-20^). The next most significant cell types were B cells (cell type #0, *p*=4.7x10^-14^) and natural killer cells (cell type #2, *p*=3.5x10^-12^). These results are consistent with longstanding understanding of pathogenic T cell involvement in multiple sclerosis, best- available treatments for multiple sclerosis, and also with more recent findings linking B cells to multiple sclerosis^52–54^. For Alzheimer’s disease, the most significant cell types were microglial (most associated cell type: #6, *p*=2.4x10^-7^). Again, these results are consistent with longstanding (>100 years ago) and newer findings. Specifically, glial alterations were reported by Alzheimer in his 1907 neuropathological description of the disease^55^, prior work established that many Alzheimer’s disease genes are preferentially expressed in microglia^30^, and newer therapeutics targeting microglia are being investigated for Alzheimer’s disease^44,56^.

### Statistical power requirements

We also sought to determine how the statistical power of individual GWAS influenced our detection of cell type associations, hypothesizing that poorly powered GWAS might not afford discovery of cell type associations. We further hypothesized that successive GWAS with increasing statistical power (as evidenced by increasing numbers of loci detected) would also afford increasing numbers of associated cell types until most of the relevant cell types were associated (or until “saturation” was afforded by adequately powered GWAS). Saturation would be indicated by a leveling off in the number of associated cell types. Thus, we re-ran our primary analysis on four schizophrenia GWAS with progressively increasing sample sizes (and statistical power), ranging from a 2011 GWAS (N=21,865) to the most recent GWAS (N=320,404)^9,19,20,57^. **Figure 5** shows results consistent with our hypotheses. The smallest GWAS revealed no significant cell type associations in our analysis, though it was adequately powered to detect five GWAS loci. By contrast, the subsequent schizophrenia GWAS afforded detection of 63, 90, and then 109 cell types, corresponding to discovery of 108, 145, and then 287 loci as reported in the primary publications^9,19,20^. Thus, we demonstrated that cell types can be linked to psychiatric phenotypes with this approach *when adequately powered GWAS are available,* and that “saturation”, of cell type discovery appears to occur at much smaller sample sizes than “saturation” of loci (predicted to saturate at >10,000 associated variants for psychiatric disorders^4,5^). The plateauing of associated cell types suggests that relevant biological features are being captured at the cell type level, via aggregation of polygenic signal into cell types.

**Figure 5.**
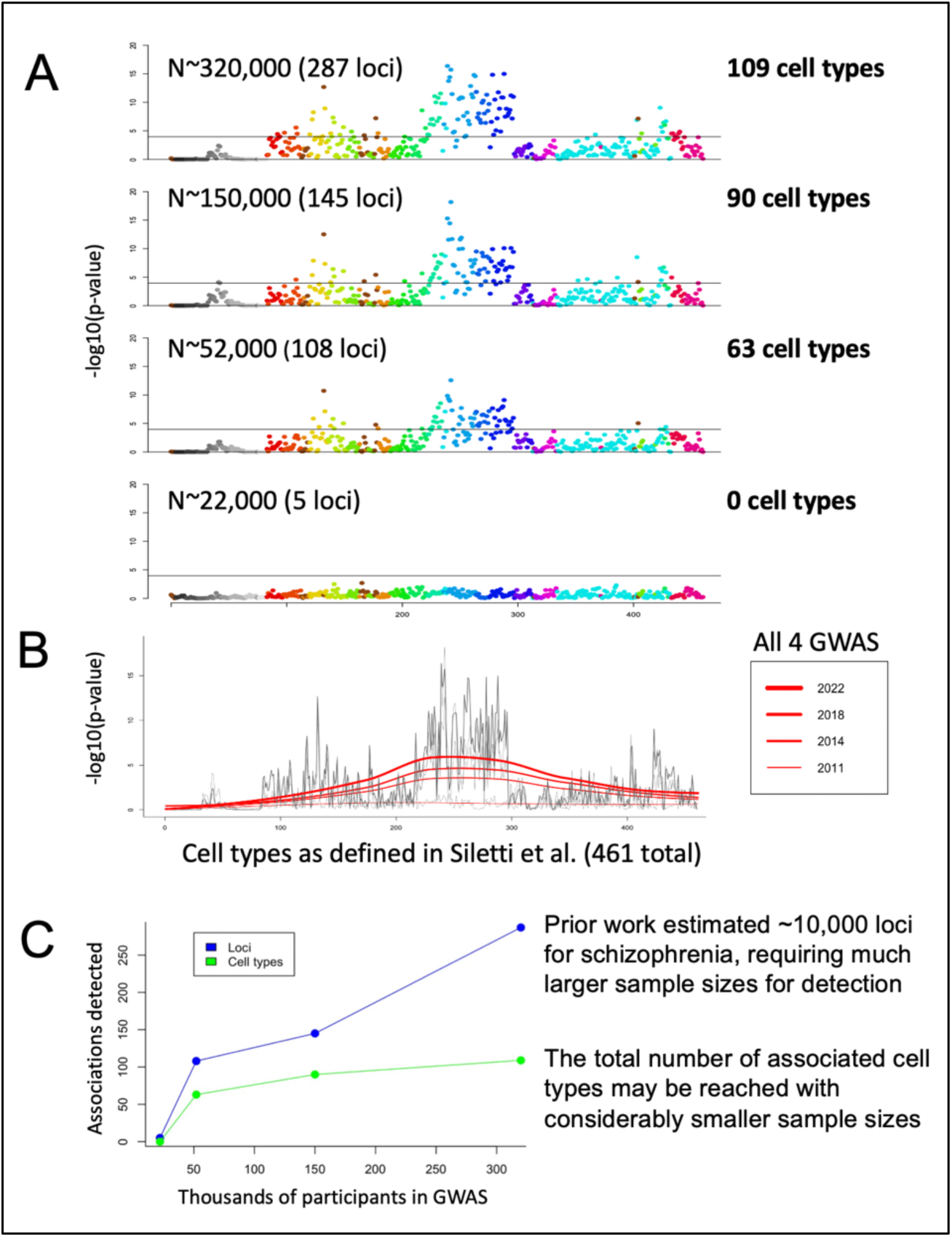
Demonstration of required statistical power of the GWAS for detecting cell type associations. **A.** Results for four successive, increasingly larger, and better powered, schizophrenia GWAS. Horizontal lines in all plots denote Bonferroni corrected statistical significance for 461 cell types (p<.0001). **B.** All four sets of results superimposed with loess lines in red. **C.** In this range of sample sizes for schizophrenia GWAS, the number of loci detected continues to increase steeply, but the number of cell types plateaus.

### Towards a cellular psychiatric taxonomy

This work demonstrates the possibility of building a taxonomy for psychiatric disorders based on quantitative evaluation of cell types. **Figure 6A** illustrates this idea for the five examined phenotypes: schizophrenia, alcohol consumed per week, sleep per night, multiple sclerosis, and Alzheimer’s disease. The proposed cellular taxonomy naturally structures future exploration, both statistical and experimental. As shown in **Figure 6B**, future statistical investigations can test for evidence consistent with schizophrenia-linked cell types acting together. For example, particular combinations of cell types act together in small local circuits, while others function together through longer-range projections. Pairs, triplets, or any other combination of cell types may be tested together for associations with schizophrenia. **Figure 6C** depicts another way that cell-type associations with schizophrenia may ultimately improve schizophrenia treatment. By examining the molecular properties of schizophrenia-associated neurons (e.g. neurotransmitter usage, receptor expression, etc.), scientists might repurpose currently available drugs, or design new drugs, to target these cell types, thus modulating specific dysfunctional brain circuits. By examining other cell types for the expression of relevant receptors, it may also be possible to predict side effects. Perhaps most excitingly, future work might even parse genetic risk for schizophrenia *at an individual level* using cell-type or circuit-level polygenic scores. The point of this endeavor is not to explain the most phenotypic variance (as is the goal of full-genome polygenic scores), but rather to predict the most vulnerable cell types for a given individual, and to accordingly tailor treatment. Note that the cost of these individualized predictions should be minimal, requiring only GWAS genotyping (<$50 USD) and analysis (extremely inexpensive after development of algorithms; i.e., ∼$150 USD and subsequently decreasing).

**Figure 6.**
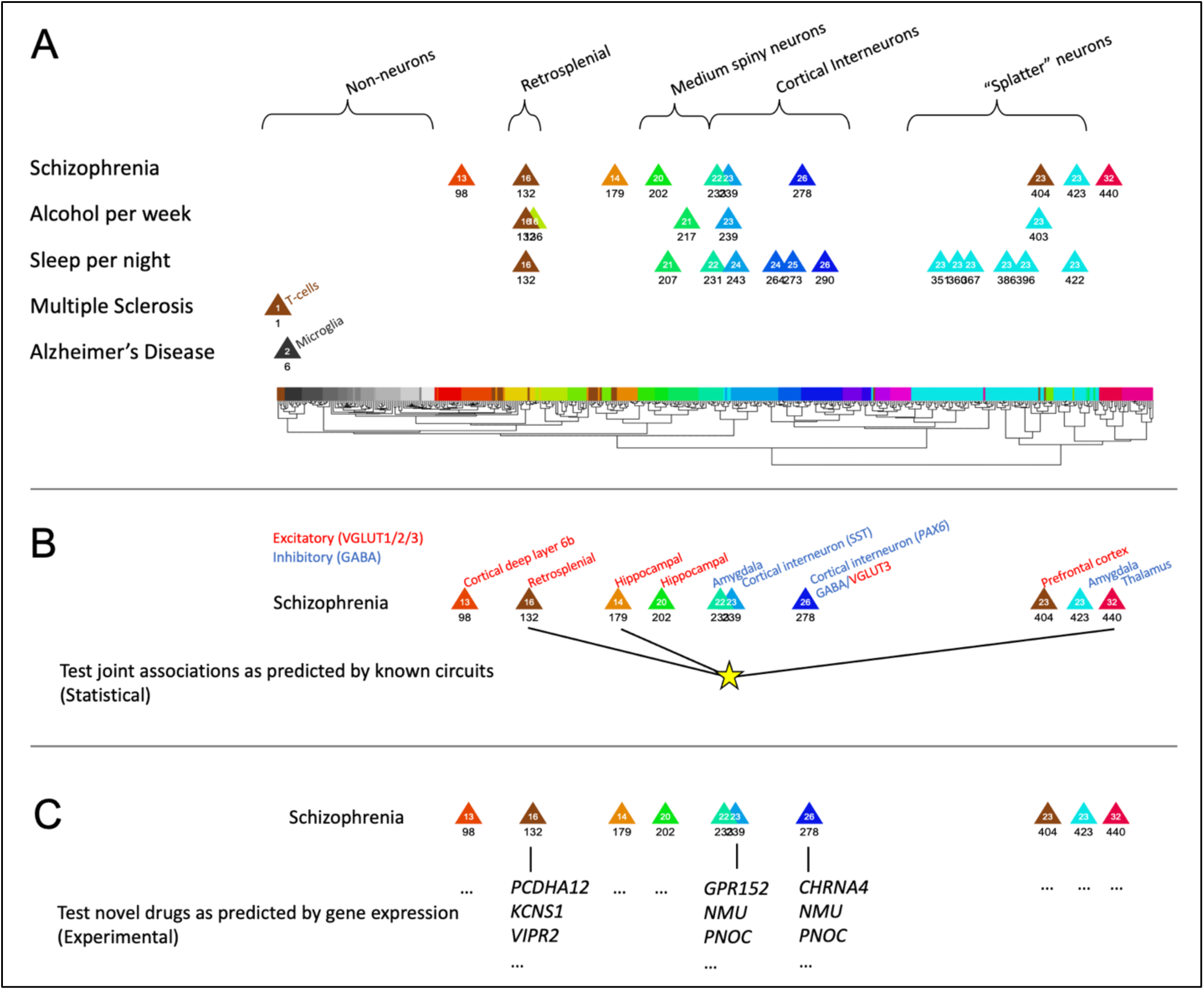
A cellular taxonomy for brain phenotypes maps shared and non-shared genetic influences in a mechanistically informative manner, and implies next steps toward deeper mechanistic understanding of schizophrenia as well as novel, personalized treatments. The 31 superclusters are color coded as in Figure 2. **A.** Significant independent cell types are depicted for the five analyzed phenotypes. Given that psychiatric disorders have shared and non-shared genetic influences; this approach may reveal the cellular (and therefore mechanistic) relevance of these shared and non-shared cell type associations. For example, shared or ‘pleiotropic’ cell types include cortical interneurons (#239) and excitatory retrosplenial cortex neurons (#132). Regarding non-shared cell types, sleep-specific associations of pons (#396) and medulla (#386) “splatter” neurons are notable.

Future work may extend to additional psychiatric disorders or subtypes of psychiatric disorders (e.g. positive, negative, and/or disorganized symptoms of schizophrenia). **B.** Statistical follow-up approaches include testing for joint association of cell types known to function together in circuits. **C.** Experimental follow up approaches include testing of drugs as predicted by gene expression in associated neuron types. Examples of predicted targets, in three schizophrenia associated cell types (#132, #239, #278), are given for illustration purposes, but data afford much more extensive predictions, for all cell types.

## Discussion

Major scientific advances often depend on newly available data obtained either in greater detail or scale, or via combined datasets capable of yielding novel insights. The vision and perseverance of leaders of the Psychiatric Genomics Consortium (PGC) enabled discovery of thousands of specific risk loci for schizophrenia and other psychiatric disorders, using unprecedented sample sizes (>1 million participants)^9,18,19,29^. Combining the PGC schizophrenia dataset with another landmark data resource, a single nucleus RNA sequencing (snRNAseq) dataset of 3,369,219 unique cells from 106 human brain regions^6^, we were able to use robust analytical approaches^26^ to discover cell types likely to be directly involved in the etiology of schizophrenia based on their expression of schizophrenia associated genes. To our knowledge, this is the first time that such a comprehensive catalog of human brain cell types has been systematically tested for associations with schizophrenia.

Cell type associations consistent with prior findings (discussed throughout the manuscript) lend confidence to our results, but novel findings can broaden our understanding of these disorders. Highlighting just two examples, we note that fear and the sense of self are two symptom domains that are clinically important for many patients with schizophrenia. Indeed, fear is one of the defining emotional features for many who suffer from schizophrenia and the amygdala is critical for fear processing. Thus, the amygdala cell types reported here (#233, #423) may be causally linked to the maladaptive fear that causes so many problems for individuals with schizophrenia. A more amorphous, but nevertheless clinically relevant and historically notable feature of schizophrenia concerns distortions in the sense of self. Consequently, it is noteworthy that we discovered a highly associated cell type (#132) in a brain region (retrosplenial cortex) that is critical for the sense of self. Specifically, recent findings showed that a particular rhythm in the retrosplenial cortex causes dissociation^58^, meaning alterations in one’s sense of self. In other words, we found that neurons in a brain region critical for an integrated sense of self are associated with schizophrenia, a disorder noted for alterations in the sense of self. Moreover, this retrosplenial neuron type is significantly associated with many psychiatric phenotypes, and thus may represent a common node of dysfunctional processing in psychiatric disorders.

Historically, classifying psychiatric disorders has proven challenging, generating enduring debate about psychiatric nosology. Our final figure depicts cell type profiles for the five phenotypes analyzed here, providing a proto-taxonomy for these phenotypes. Here we suggest that **cell typologies may prove the right level of analysis for a stable taxonomy of brain disorders; making available the appropriate measurements and organizing principles.** What remains to be determined is the relevance of developmental versus adult cell types to different disorders, the identifiability of subtypes of disorders with this approach, and the eventual impact of this research on classification of psychiatric disorders.

A cellular classification approach might be further refined using GWAS of different subtypes of psychiatric disorders. Appealingly, the long-postulated heterogeneity within psychiatric disorders (e.g., the ‘schizophrenias’) may be wrangled into comprehensibility with these approaches. Different cell type associations might be identified, for example, for major symptom domains of schizophrenia, including positive, negative, and disorganized symptoms. Further, by specifying molecular properties of etiologically relevant cell types for different disorders (e.g. receptors and neurotransmitters) this classification system should naturally nominate treatment targets, as well as relevant therapeutics. By analogy, quantifying and organizing electrons and protons allowed classification of elements into the periodic table and thus generated novel predictions about elemental interactions. A cell-typology for psychiatric disorders may similarly afford novel predictions about which medications and treatments are most likely to be effective for each disorder. One such approach involves developing polygenic risk scores for cell types or circuits. Individual level metrics of genetic risk, polygenic risk scores, are already available for multiple psychiatric disorders. Typically, polygenic scores use variants from across the genome to maximize accurate prediction of risk. To determine the best medications for individuals, however, cell-type restricted polygenic scores may eventually prove more useful.

## Limitations

There are important limitations to consider regarding this approach and currently available datasets. First, this approach cannot discover cellular abnormalities resulting exclusively from environmental influences. Conversely, this “limitation” also implies that reported cellular associations do not result from environmental risk factors, treatment, or consequences of living with schizophrenia. Second, although these datasets represent landmark achievements in GWAS and transcriptomically-derived cell types, they have important limitations, which diminish the ultimate power of this approach. Briefly, this snRNAseq dataset^6^ did not include all brain regions, the female sex, or non-European ancestry individuals. Specific brain regions not sampled in Siletti et al. which may be particularly important for schizophrenia include the dorsolateral prefrontal cortex (DLPFC Broadmann area 9) and the ventral tegmental area of the midbrain (VTA). Further, since this snRNAseq dataset came from adult humans, an important next step will be conducting the same analyses using developmental datasets. For example, another dataset from the Linnarsson group^59^ includes human brains from post-conception weeks 5-14, a time period critical for neurogenesis and potentially of importance for psychiatric disorders.

The statistical methods used here were selected for their robustness to false positives and prior use with transcriptomic datasets, but such models may be further optimized, for example via inclusion of data from exome sequencing studies of schizophrenia. Further, we used a straightforward method of linking genetic variants to genes based only on proximity, and thus longer-range effects of variants on genes were omitted from our analyses. Finally, to the extent that the central assumption of our analysis method – that cell types that make greater relative use of schizophrenia-associated genes than other cell types – is incorrect, cell type associations reported may be incorrect.

In conclusion, the results reported here are possible thanks to a combination of natural endowment (the genome as a catalog of biological elements) and human technology (i.e., modern advances which allow us to query the genome – in both RNA and DNA forms – in great detail). Together, these results provide a new framework for understanding the cellular basis of psychiatric disorders. Given the detailed molecular information available about each cell type (e.g. receptors, neurotransmitters, neuropeptides), this work also provides a roadmap toward novel therapeutics, better use of existing medications, and truly personalized medicine.

## METHODS

### Selecting comparison phenotypes

In addition to schizophrenia, we selected four comparison phenotypes (alcohol consumed per week, sleep duration per night, multiple sclerosis, and Alzheimer’s Disease). These phenotypes met the following criteria: 1) they were polygenic, and 2) well powered GWAS of those phenotypes were available (both conditions are required for this method). Regarding psychiatric comparison phenotypes, we wanted to mitigate concerns about the reliability of psychiatric diagnoses^v^, so we chose alcohol consumed per week and sleep duration per night because they are readily quantifiable (albeit noisily). The two neurological phenotypes (multiple sclerosis and Alzheimer’s disease) were selected because have considerably different etiology from psychiatric disorders^60^, and consequently they provided an opportunity to observe greater contrasts among this set of five brain-related phenotypes. Genome-wide association studies (GWAS) results used here were from the best powered GWAS of these phenotypes available at the time of analysis (Nov 2022-July 2023). The relevant sample sizes and source publications were as follows: schizophrenia^9^ (N=320,404), alcohol per week (N=2,428,851), sleep duration per night^11^(N=446,118), multiple sclerosis^30^ (N=41,505), and Alzheimer’s disease^31^ (N=788,989). Given that well powered GWAS are required for these analyses, we used only European ancestry samples since adequately powered GWAS are not available for any other ancestry, for all these phenotypes.

### Gene expression data

The gene expression data from Siletti et al. is publicly available. Briefly, Siletti et al. sampled 3,369,219 nuclei from 106 different dissections in three postmortem brain donors using high-throughput 10X Chromium single-nucleus RNA sequencing. The nuclei were then computationally clustered into 31 superclusters and 461 cluster. The 461 clusters are referred to as “cell types” in our paper. For data processing, we first applied a transformation to the single-cell expression data to compress the scale and reduce outliers (as is typical for such data, here we used *ln*(1 + *x*)), and obtained the mean transformed expression for each gene in each cell type. We only kept protein-coding genes from the National Center for Biotechnology Information (NCBI) and removed unexpressed genes, genes with non-unique names, and genes within the MHC region (chromosome 6 base positions 25,000,000- 34,000,000). For MAGMA use, we mapped Ensemble IDs to Entrez gene IDs with the Genome-Wide Association for Human package (“org.Hs.eg.db”) in Bioconductor (v3.12.0).

Genes without unique Ensemble-Entrez mappings were removed. A metric of gene expression specificity was then calculated by dividing the transformed expression of a gene in a cell type by the sum of the transformed expression of that gene across all cell types, yielding a value between 0 and 1 that characterizes the extent to which a particular gene is expressed in a particular cell type. For example, in hippocampal neuron type 202, 12% of the total transformed gene expression of the *GUCA2A* gene is in this cell type, so the specificity score for *GUCA2A* in cell type 202 is 0.12. If a hypothetical gene were completely evenly expressed across all cell types, then the specificity score for that gene, in all cell types, would be 1/461 = 0.0022.

### MAGMA overview

MAGMA (v1.10) is software designed for gene and gene set analysis of GWAS data, and it has been extensively tested to ensure appropriate control of type I errors and adjustment for potentially confounding variables^26,61^. MAGMA uses a regression framework and employs a two-stage procedure to test for associations, first calculating gene level p-values and then using those gene level p-values to compute p-values for collections of genes. Collections of genes can either be analyzed as gene sets (using binary coding of genes that are in or out of the set) or as ‘gene properties’ meaning quantitative values assigned to all genes, as we have here (i.e., specificity values for each gene, in each cell type). We employed the gene property analysis rather than arbitrarily imposing a threshold on the specificity scores to define a gene sets for each cell type. We also used the optional third stage, conditional analysis, to specify likely independent associations among all significant associations.

### MAGMA gene level analysis

We first used MAGMA to map each SNP to a gene if the SNP was located within 35 kilobases (kb) upstream to 10 kb downstream of that gene. We then used MAGMA’s SNP-wise mean (snp-wise=mean) model to conduct gene analysis while adjusting for linkage disequilibrium (LD). LD data was from the European ancestry panel of 1000 Genomes phase 3^62^. In gene analysis, the test statistic of a gene was calculated as the sum of squared SNP z-statistics, where z-statistics were the probit transformation of SNP p-values from GWAS. Because the test statistic for each gene followed a mixture of independent χ^2^_1_ distributions under the null hypothesis, we calculated gene p-values (each representing the association between a phenotype and a gene) accordingly.

### MAGMA gene property analysis

MAGMA’s gene property analysis represents the association that a gene has with a given phenotype as a z-score *Z*_*g*_ = probit(1 − *Pg*) where *P*_*g*_ is the p-value of a given gene from the gene analysis step in MAGMA. Per MAGMA default, we truncated z-scores that were 3 standard deviations below or 6 standard deviations above the mean to prevent outliers from biasing analysis results. We then conducted the gene property analysis via a linear regression model *Z* = β_0_ + *P*_*c*_β_1_ + *C*β_1_ + ε (*eq*. 1), where *Z* is the aforementioned z-scores of each gene, *P*_%_ is the specificity of each gene in a given cell *c*, *C* represents the covariates, and ε is modeled as a multivariate normal accounting for the LD between genes. Per MAGMA default, specificity values were truncated if they were 5 standard deviations from the mean. In our analysis, covariates were gene size, gene density, sample size, inverse mean minor allele count, and their log values. Lastly, we conducted a one-directional test of the coefficient β_!_as a test of the association of each cell type with each phenotype. For each phenotype (e.g., schizophrenia), this analysis was run 461 times (i.e., once for each cell type).

### MAGMA conditional analysis

To specify likely independent signals from among all significant results (i.e. all significant cell types for each phenotype), we conducted pairwise conditional analyses using MAGMA. Here, we used the linear regression model 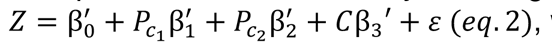, which is the same as the one from gene property analysis except that the model here includes two cell types of interest. We then conducted forward stepwise selection as detailed in Watanabe et al.^23^ to arrive at a set of “independent significant” cell types. For cell type *c*_1_ and *c*_2_, let us denote the p-value associated with β^1^′ in *eq*. 2 be *p*_*c*_1_,*c*_2__ and the one associated with β′_2_ be *p*_*c*_2_,*c*_2__. We also denote the marginal p-values associated with the respective cell types from gene property analysis be *p*_*c*_2_,*c*_1__ and *p*_*c*_2__. We define proportional significance, which portrays the remaining significance of a cell type *c*_1_ after conditioning on *c*_2_, as *PS*_*c*_1_, *c*_2__ such that 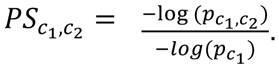. In forward stepwise selection, the set of independent significant cell types (denoted as *S*) initially only contained the most marginally significant cell type. The next most significant cell type *c* was added to the set *S* in succession only if it satisfied two scenarios: First, if both *PS*_*c,s*_, *PS*_*s,c*_ ≥ 0.8 for all *s* ∈ *S*, the associations of cell type *c* and any *s* ∈ *S* with a given phenotype were considered independent. Second, if 0.5 ≤ *PS*_%,3_ < 0.8, 0.5 ≤ *PS*_3,%_ < 0.8, and *p*_*c*_ ≤ 0.05 for all *s* ∈ *S*, the associations of cell type *c* and any *s* ∈ *S* were only partially explained by each other, while the majority of the signals were independent. Cell types not included in the set *S* can still play an important role in the etiology of a phenotype; however, the selection procedure excluded them because their association cannot be distinguished from the association of cell types in *S*. Note that in some rare cases, cell type with a lower marginal significance can have a higher conditional significance. When *PS*_*c1, c2*_ < 0.2 yet *PS*_*C2,c1*_ ≥ 0.2 for cell type *c*_1_ and *c*_2_ where *p*_*c1*_ < *p*_*c2*_, the order of the selection process was reversed for the two cell types.

### Considerations regarding anatomical annotations

Throughout this manuscript, we refer to the anatomical locations of cell types. These locations were reported in Siletti et al., whose procedures involved sampling from of 106 separate human brain regions (“dissections”), and to which each of the 3,369,219 cells in their study can be traced. Note, however, that it is rare for any of the 461 cell types to be exclusively from any one dissection or brain region. Indeed, one goal of such transcriptomic surveys is to reveal the ways in which transcriptomically similar cells are found in different areas of the brain.

**Supplementary** Figure 5 summarizes information about the dissections contributing to each cell type. For example, interneurons, which are inhibitory neurons found widely across the cortex (cell types #236-296 in Siletti et al.) derive from dozens of cortical (and even some subcortical) regions. Accordingly, the single dissection contributing the most cells to any of the interneuron clusters typically accounts for <10% of cells in the cluster. Conversely, cell type #202 is from just two dissections, both in the hippocampus. For brevity in the manuscript, we refer to cell types as “hippocampal” or “amygdala”, etc. when cell types derive primarily from that brain region (i.e. >50%, but typically much higher). Precise information about the dissections accounting cells in each cell type is given in **Supplementary Table 1**, which provides the total number of cells in each cluster and the percentage of cells in that cluster that came from each contributing dissection.

### Note about cell types / cell nomenclature

Nomenclature for cell types will continue to evolve as the naming and categorization of cell types continues. For clarity, we have not addressed the issue of continuous gradients of difference across cell types, but this will be an important area of investigation for future studies.

## Supporting information

Supplementary Tables 1 through 5

## Acknowledgements

This work was supported by the Jaswa Innovator Award to LD and by the National Institute of Mental Health (NIMH) to LD (R01 MH123486 & R21 MH125358).

## Author contributions

L.D. designed the experiment, reviewed all data and analyses, drafted the manuscript, made the figures; T.L., M.S, W.L, H.S., N.S, S.V, & H.S. analyzed the data; J.Y., G.W., J.B., L.T., B.S.P., B.K., K.D, & W.J.G provided essential expertise; all authors edited and approved the manuscript.

## Materials & Correspondence

Correspondence should be directed to L.D.

## Data availability

All datasets used here are publicly available from the primary publications as noted throughout the manuscript.

## SUPPLEMENTARY FIGURES

**Supplementary Figure 1.**
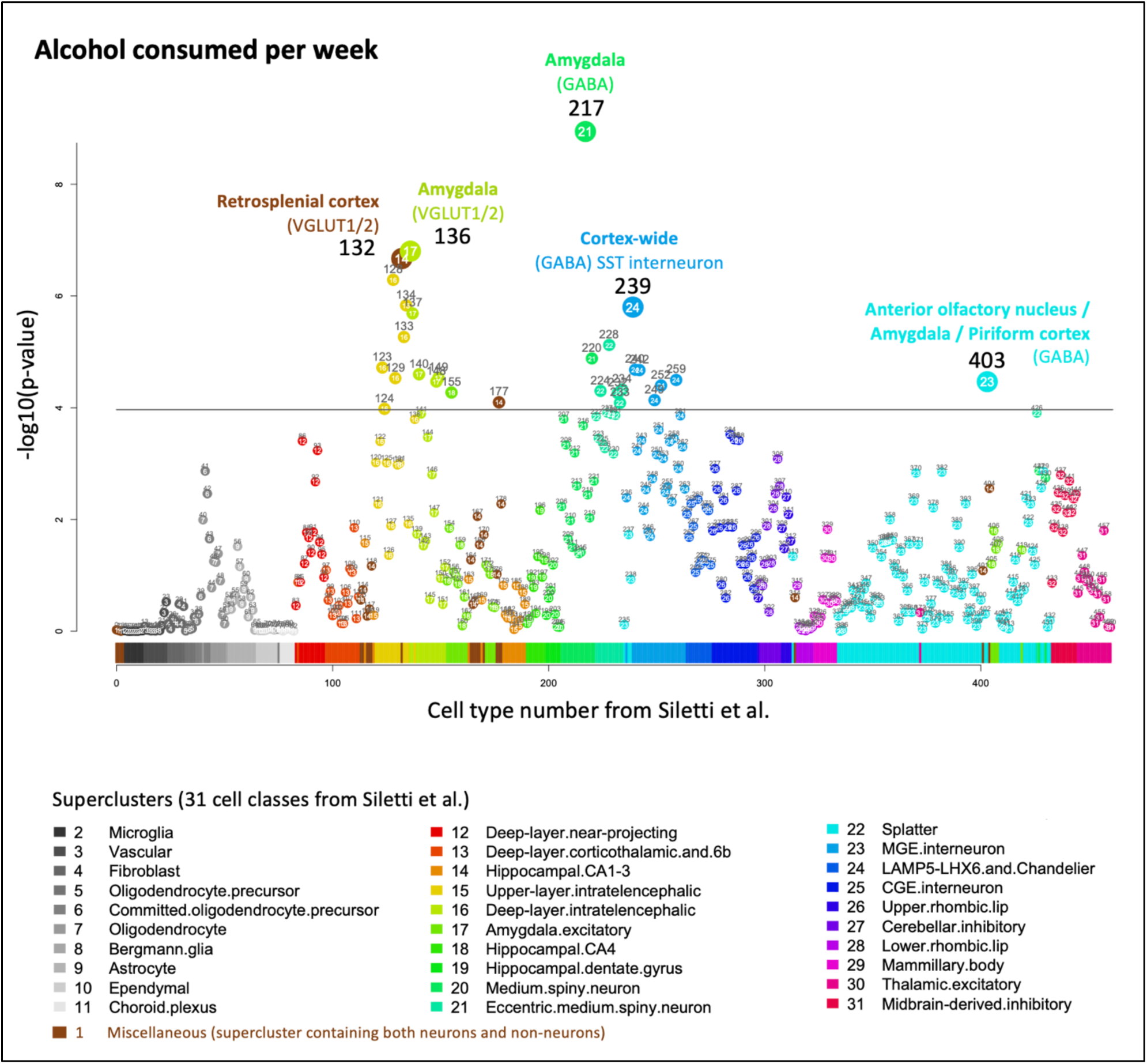
Cell type associations for alcoholic drinks consumed per week.

**Supplementary Figure 2.**
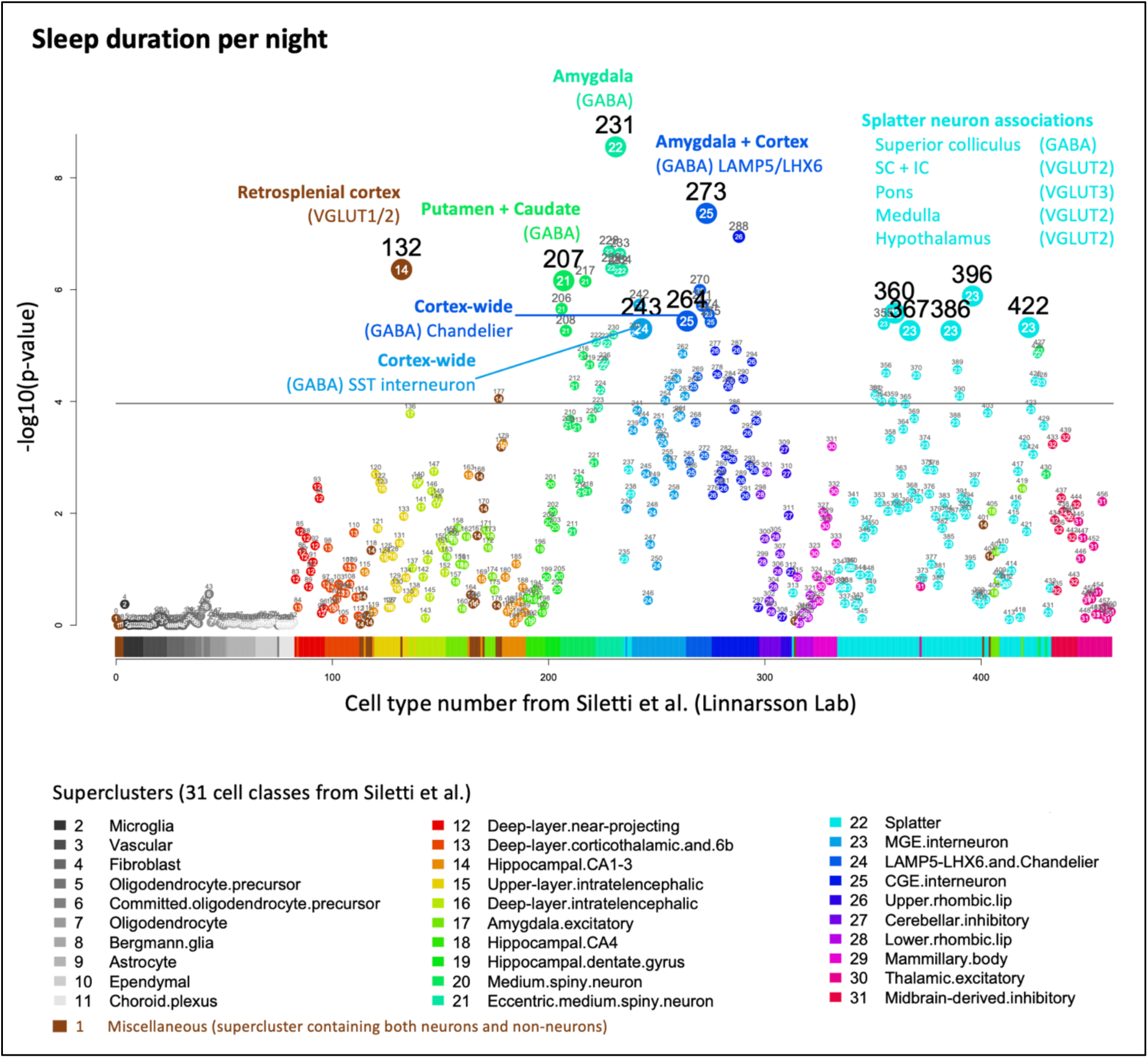
Cell type associations for sleep duration per night.

**Supplementary Figure 3.**
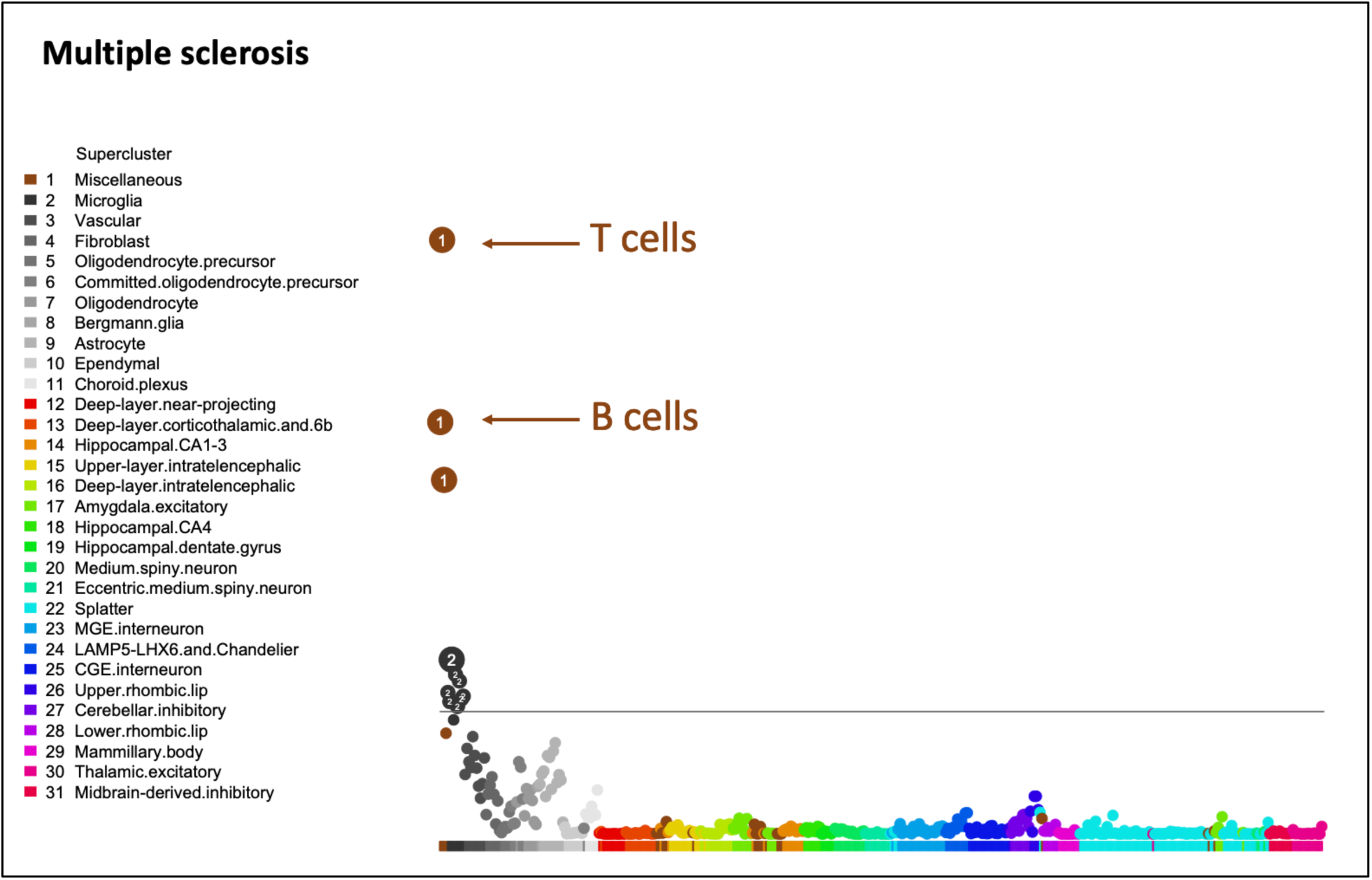
Cell type associations for multiple sclerosis

**Supplementary Figure 4.**
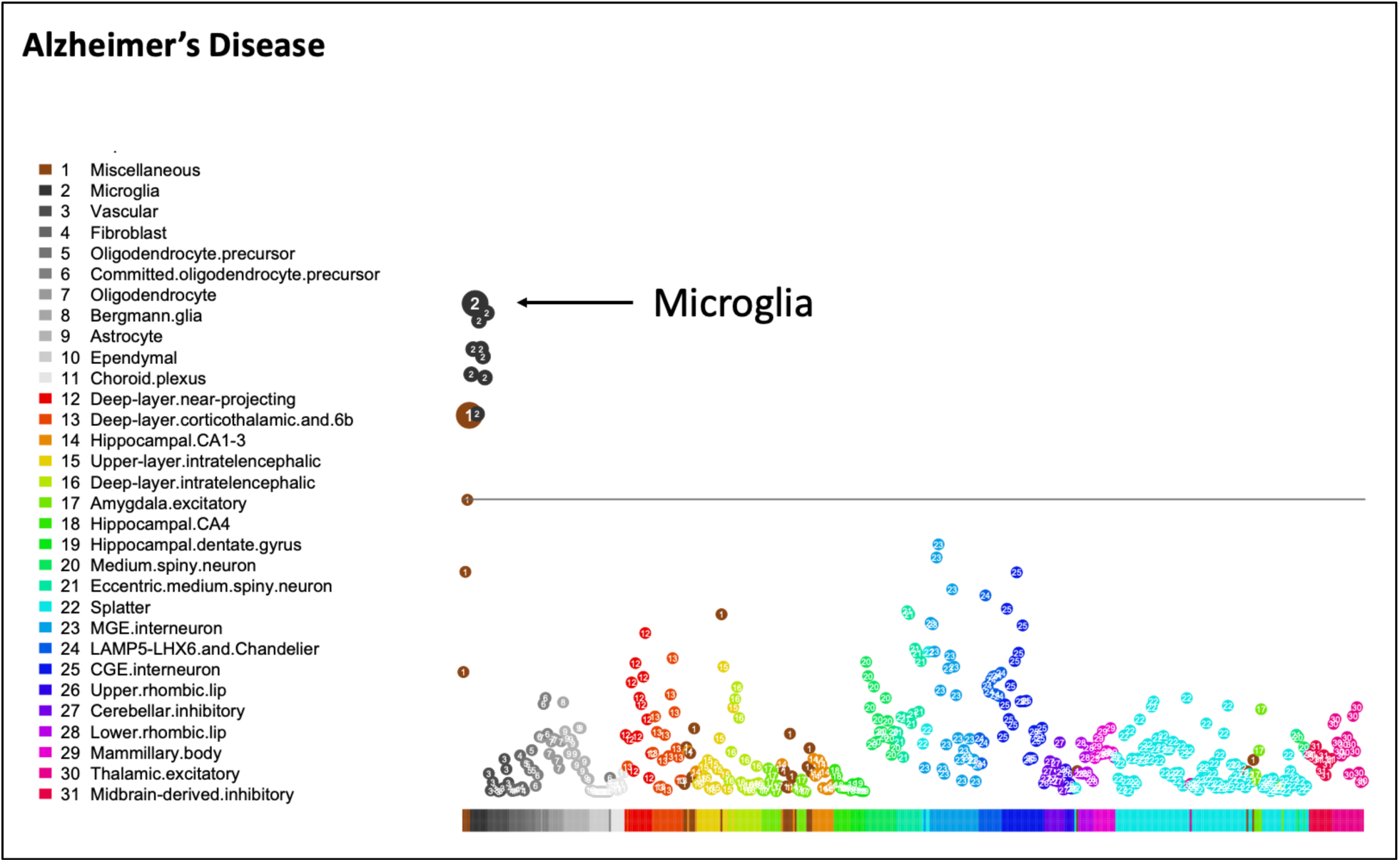
Cell type associations for Alzheimer’s disease

**Supplemental Figure 5.**
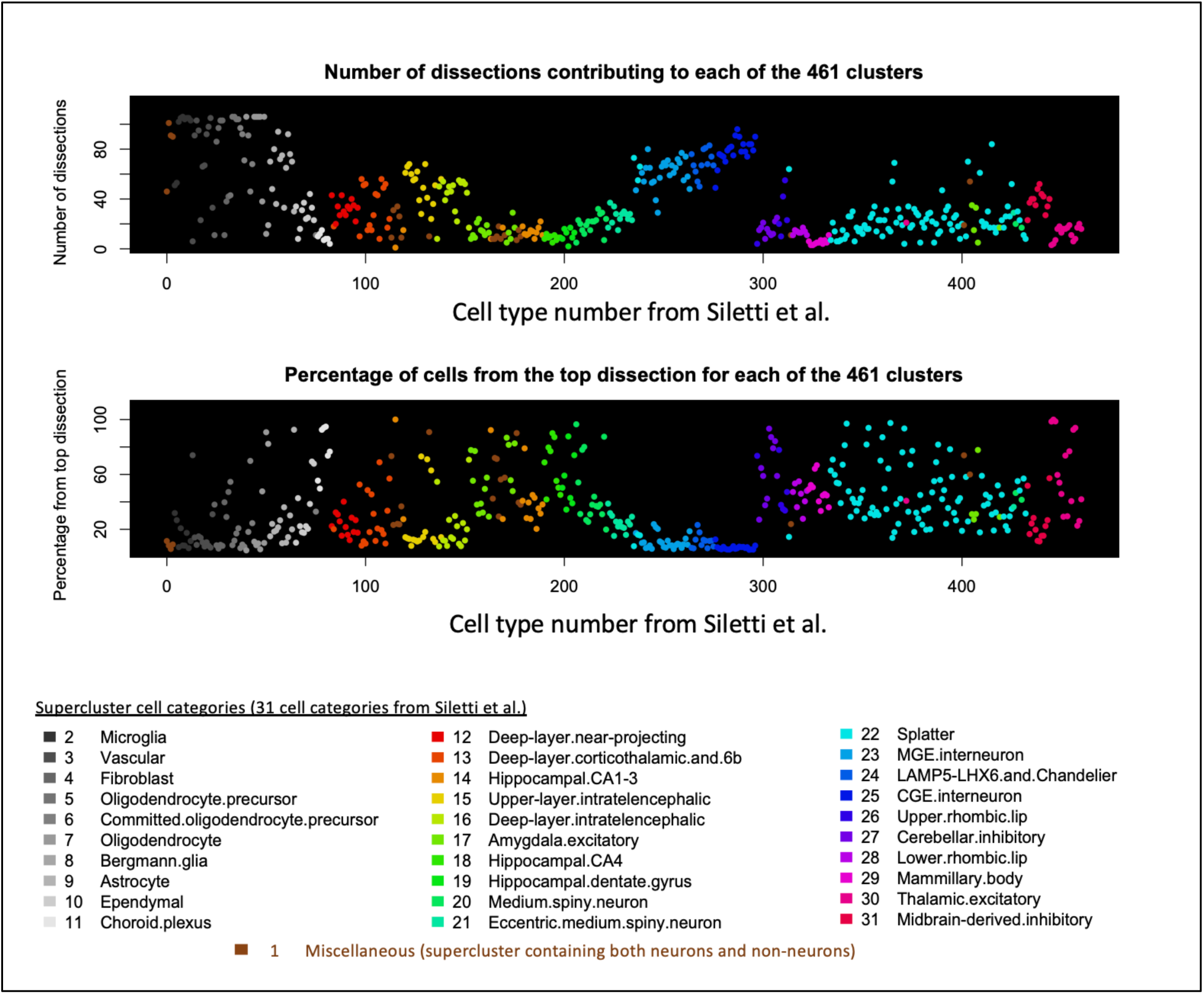
I**n**formation **about the number of dissections contributing to each of the 461 cell types.** For example, interneurons (cell types #236-296), shown in blue shades, are among the cell types that are found in the largest number of dissections (top) and no single dissection accounted for >25% of any interneuron type, and for most the top contributing dissection accounted for <10% of the cells in that cell type (bottom). The number of contributing dissections for these widely-distributed interneuron cell types was 56-84 out of 106 possible dissections.

## Footnotes

i “scRNAseq” when single cells are used instead of snRNAseq when single nuclei within cells are used.

ii We also examined cell type #242 because it was nearly as significant as the top cell type (#239). This second-most significant cell type (#242) was also associated with other psychiatric phenotypes, was larger than #239 (i.e., encompassed ∼3x more cells in SileF et al), and it was the most significant cell type in our analysis of the 2018 schizophrenia GWAS. Thus, we thought #242 was also a strong candidate for psychiatric relevance.

iii Note that we also found medium spiny neurons from the striatum to be associated with schizophrenia (#222 and #224).

iv Commi%ed oligodendrocyte precursors (cOPC) were the most significant non-neuronal cell types for psychiatric phenotypes. Note that Bonferroni correction for 461 cell types is overly stringent, and we are not arguing that glia play no role in psychiatric phenotypes.

v LimitaQons notwithstanding, psychiatric diagnoses are moderately reliably diagnosed across Qme and raters. Further, unbiased clustering approaches applied to symptom level and geneQc data support the uQlity and validity of such diagnoses.

